# Magneto-optical diagnosis of symptomatic malaria in Papua New Guinea

**DOI:** 10.1101/2020.05.14.20101543

**Authors:** Leandra Arndt, Tamarah Koleala, Ágnes Orbán, Clemencia Ibam, Lincoln Timinao, Lina Lorry, Ádám Butykai, Peter Kaman, Andrea P. Molnár, Stephan Krohns, Elma Nate, István Kucsera, Erika Orosz, Brioni Moore, Leanne J Robinson, Moses Laman, István Kézsmárki, Stephan Karl

## Abstract

Improved methods for malaria diagnosis are urgently needed. Here, we evaluated a novel diagnostic test named rotating-crystal magneto-optical detection (RMOD) in 964 suspected malaria patients in Papua New Guinea. RMOD tests can be obtained within minutes and at low cost. Capillary blood samples were also subjected to rapid diagnostic tests, expert light microscopy and polymerase chain reaction to systematically evaluate the capability of RMOD to detect infections. Compared to light microscopy, as the gold standard, RMOD exhibited 82% sensitivity and 84% specificity to detect any malaria infection. This increased to 87% sensitivity and 88% specificity for *Plasmodium vivax*, indicating that RMOD could be useful in *P. vivax* dominated elimination settings. Parasite density correlated well with the quantitative magneto-optical signal. Importantly, residual hemozoin present in malaria negative patients was also detectable by RMOD, indicating its ability to detect previous infections, which can be exploited to reveal transmission hotspots in low-transmission settings.

## INTRODUCTION

Humans have suffered from malaria for thousands of years and still, hundreds of millions of people are infected each year. Nowadays, malaria also places a significant social and economic burden on tropical developing countries, further undermining potential for growth.[1] Development of rapid, easy-to-use and low-cost malaria diagnostic methods, with high sensitivity and specificity, remains an urgent priority in tropical diseases research.[2, 3] Currently available methods include the inspection of blood smears using light microscopy (LM), rapid diagnostic tests (RDTs) and molecular methods, such as polymerase chain reaction (PCR). These techniques rely on different diagnostic targets, namely the direct observation of infected red blood cells (LM), detection of parasite antigens (RDT) or DNA (PCR).

Researchers have also been fascinated by the magnetic properties of malaria infected red blood cells for a long time since using an inherent and unique physical property such as malaria parasite-induced red cell magnetism, may enable rapid and easy diagnosis at low cost. The increased magnetic susceptibility of red blood cells infected with *Plasmodium* parasites is a striking, and well described biophysical phenomenon arising from the metabolization of hemoglobin.[4–6] Normally, oxygen-bound hemoglobin is a diamagnetic substance with a magnetic susceptibility close to that of water.[7] During infection of red blood cells, *Plasmodium* parasites break down hemoglobin in their digestive vacuoles and heme molecules liberated in the process are assembled into iron-containing organic crystallites called hemozoin.[8–12] These hemozoin crystals are one of the most distinguishing features of malaria infection in peripheral blood and played a vital role in identifying *Plasmodium* parasites as the cause of malaria and the mosquito as the agent of transmission in the late 19th century.[13, 14] During the process of hemozoin formation, hemoglobin iron is oxidised and concentrated to make up about 10% of the mass of the newly formed hemozoin crystals, resulting in an overall paramagnetic behaviour of hemozoin.[4] As such, hemozoin is an intrinsic biomagnetic marker of infection with *Plasmodium*. While the hemozoin crystals are not excreted naturally by the infected cells until cell rupture, they can easily be made accessible for diagnostic purposes by lysing blood.

Several approaches to exploit these magnetic properties for diagnostic purposes have been proposed [15–27] and some of the techniques developed, showed promise under laboratory conditions. However, evaluations of magnet-based tests in larger in-field trials are rare and have, so far, been disillusioning.[16–18, 28, 29]

Over recent years, we developed a novel diagnostic technique, named rotating-crystal magneto-optical detection (RMOD), targeting the aforementioned hemozoin material. We demonstrated that hemozoin can be detected in lysed blood with very high sensitivity in the low ng/*μ*L range using RMOD.[27] Measurements on parasite cultures indicated that RMOD had a limit of detection of *∼* 10 parasites*/μ*L of blood in samples spiked with *P. falciparum*.[27] These promising results were further supported by studies on *P. berghei* and *P. yoelii*-infected mice.[24, 25] On an operational level, RMOD is promising as it can be conducted after minimal training and provide test results within minutes. From a funding perspective, since no expensive reagents are used, per sample measurement cost is very low.

The dynamics of hemozoin accumulation and clearance in natural malaria infections in the human body are more complex than what can be mimicked in model systems, and intermediate redistribution and final fate of hemozoin during and following infections is not well understood.[35] Many mechanisms and intricacies of parasite biology and human response to infection may collectively determine the actual quantity of hemozoin in peripheral blood. For example, infected red blood cells are cleared mostly in the liver and spleen, and as a consequence these organs become loaded with hemozoin.[36, 37] Leukocytes phagocytose infected red blood cells and ingest hemozoin.[38, 39] Late parasite stages of *P. falciparum* containing large quantities of hemozoin sequester in the capillaries,[40] whereas a higher proportion of late stages of *P. vivax* keep circulating in peripheral blood. Gametocytes of *P. falciparum* contain large amounts of hemozoin and circulate for an extended period of time after an infection has been cleared or treated.[41, 42] While many aspects of hemozoin clearance and redistribution in the human body are yet to be elucidated, there is solid evidence for long-term persistence of hemozoin in body tissues of people living in endemic areas.[37]

In order to account for these complexities, which are potentially relevant to hemozoin-based malaria diagnosis, we conducted a detailed evaluation of RMOD on almost 1000 suspected malaria cases in Madang, Papua New Guinea (PNG). PNG has a complex malaria epidemiology and the study area exhibits high transmission intensities for both, *P. falciparum* and *P. vivax*.[43] Using a RMOD prototype device similar to that described in our previous studies,[24–27] we systematically compared RMOD performance to conventional diagnostic techniques, namely expert LM, RDT and PCR and found that magneto-optical hemozoin detection is a competitive approach for clinical and in-field malaria diagnosis. Fig. 1 provides an overview of the study site and population (Fig.1A); a brief comparison of conventional diagnostic techniques together with RMOD (Fig.1B), and a schematic illustration of the RMOD measurement principle (Fig. 1C – E).

**FIG. 1.**
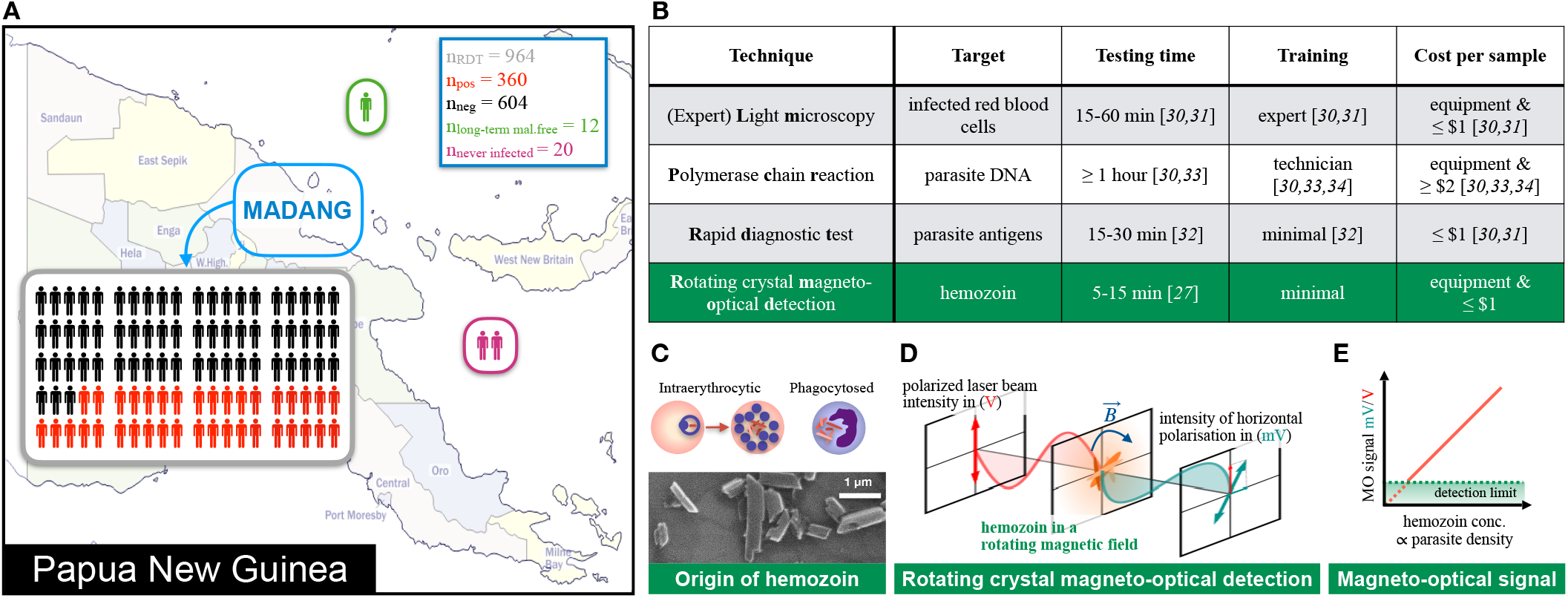
Study overview, comparison of existing malaria diagnostic techniques to RMOD and working principle of RMOD. **A** Overview of the study population in Madang, Papua New Guinea. A total of 964 suspected malaria cases were enrolled of whom 360 were found positive by expert LM. Samples from 32 malaria naïve and long term malaria free patients were also included. B Comparison of existing diagnostic techniques with RMOD in terms of their target, approximate testing time, level of training and cost per sample.[27, 30–34] C Hemozoin crystals formed by *Plasmodium falciparum*, as imaged by scanning electron microscopy. In peripheral blood, hemozoin can be present inside infected erythrocytes or phagocytosed in leukocytes. **D** RMOD principle. Polarization of the incoming laser beam is tilted due to the linear dichroism of hemozoin crystals. The rotating magnet drives a synchronous rotation of the crystals in lysed blood, which leads to a periodic tilting of the polarization and, in turn, a periodic modulation of the intensity of the outgoing laser beam. **E** The ratio of the modulated (ac) intensity and the mean (dc) intensity provides the magneto-optical (MO) signal in mV/V, which is highly-sensitive quantitative measure of hemozoin concentration and, thus, proportional to the parasite density.

## RESULTS

### Study population and infection data

A total of 964 suspected malaria patients were enrolled into the study. All of them had an RDT result, while 951 had a LM result and a PCR result. The overall properties of the patient population and the malaria diagnosis and parasitological results are given in Table 1. Overall, based on LM diagnosis 34% of patients were positive for any malaria infection, as compared to 37% and 35% by RDT and PCR methods, respectively.

**TABLE I.**
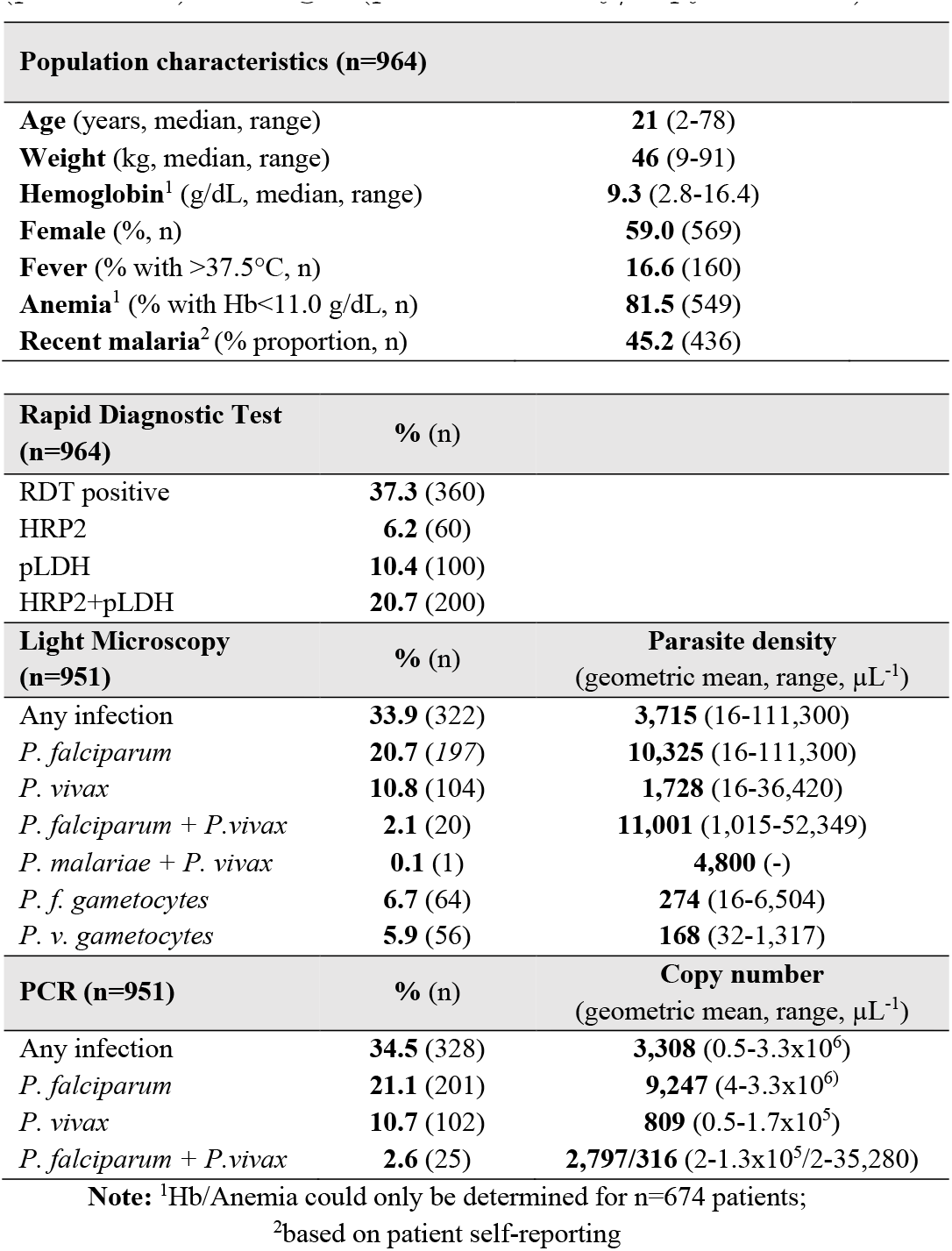
**Study population and infection data.** Characteristics of the study population (*n* = 964). Bold numbers stand for percentages or medians, while numbers in parentheses indicate ranges or number of patients. For RDT, LM and PCR results, the center column shows the percentage of positive individuals (prevalence) for the respective diagnostic method and type of infection. For LM and PCR, the right column shows parasite density (LM) and gene copy numbers (PCR) for positive individuals. Bold numbers are percentages (prevalence) or geometric means (parasite density/copy numbers). Numbers in parentheses are number of patients (prevalence) or ranges (parasite density/copy numbers).

Among the three reference methods used in this study, RDT was applied at enrolment, while we had originally planned to use Plasmodium species-specific qPCR as the main reference method. However, the considerable number of qPCR negative, yet expert LM positive samples (47 *P.f*., 35 *P.v*.) lead us to conclude that the species-specific qPCR in this study may not have performed optimally. Thus, expert LM was used as the main reference method, which is the standard recommended by WHO. Yet, the PCR results are still considered for comparison.

### Comparison of conventional diagnostic methods

When LM was used as the reference standard for comparison with RDT and PCR methods, RDT exhibited a sensitivity of 86.6% and specificity of 87.8%, while PCR showed a sensitivity of 88.9% and specificity of 80.1%. A table showing the commonly used measures of agreement (sensitivity, specificity, predictive values and Cohen’s-*κ*) is included as Supporting Information Table S1.

For the quantitative methods LM and PCR, parasite density and PCR-based gene copy number correlated well (Spearman Rank Correlation: *P. falciparum R*2 = 0.79; *p* < 0.0001; *P. vivax*: *R*2 = 0.71; *p* < 0.0001). A correlation plot is shown in Supporting Information Fig. S1.

### RMOD results

The median overall magneto-optical (MO) signals for LM positive and LM negative samples were 22.6 mV/V and 1.7 mV/V, respectively. For the RDT positive and negative samples the median MO signals were 15.0 mV/V and 1.7 mV/V, respectively. For PCR positive and negative samples the median MO signals were 7.5 mV/V and 1.7 mV/V, respectively. For all three methods, the differences in the median MO signals for positive and negative samples were highly statistically significant (Mann-Whitney test *p*-values < 0.0001). A detailed summary of the RMOD results compared to the other methods (LM, RDT, PCR) is shown in Fig. 2.

**FIG. 2.**
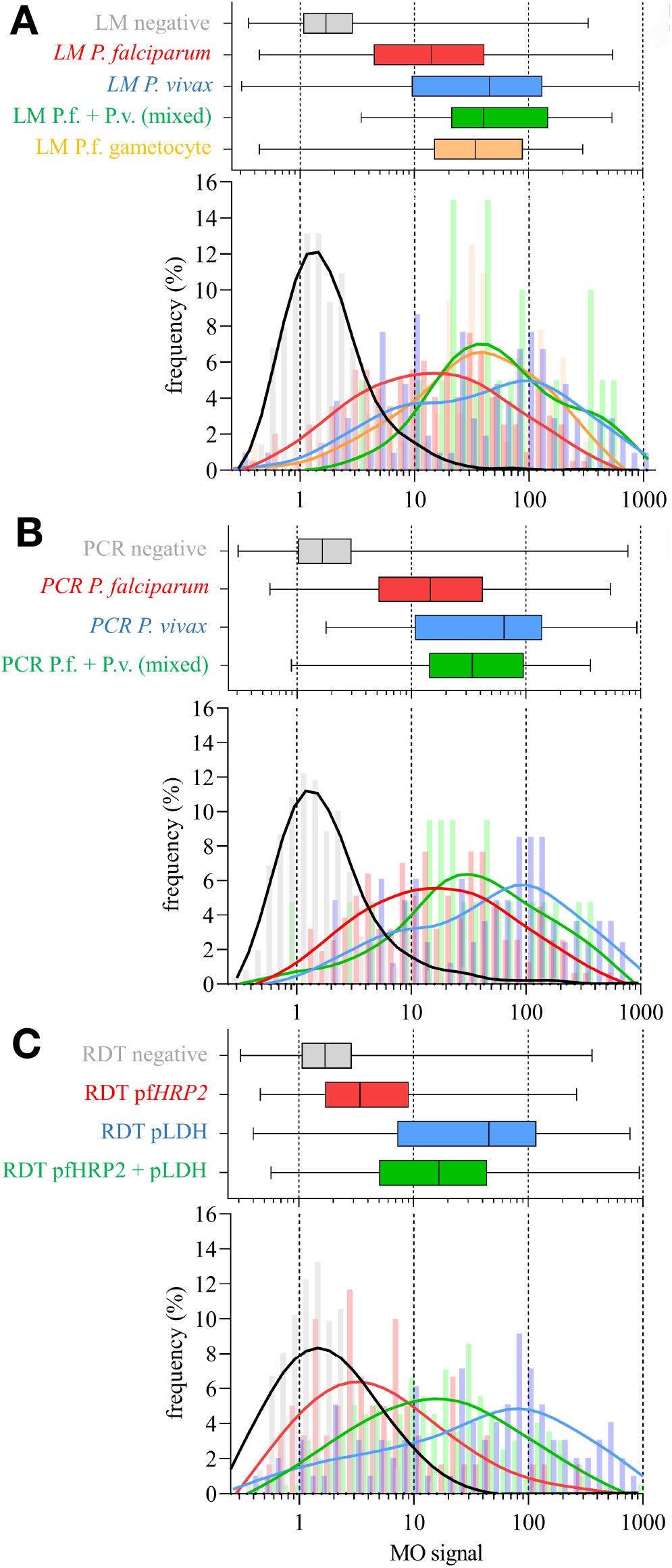
RMOD data in comparison to different reference methods. **A** expert LM, **B** PCR and C RDT. The panels are subdivided into box-and-whisker plots (top) and histograms (bottom). The box-and-whisker plots show median, interquartile range (IQR) and range of the MO signals for the respective diagnostic results. The histograms show the raw distribution of MO signal data (bars) and a smoothened line resulting from applying generalised additive models of the MO signal frequency distributions for the respective diagnostic results. All panels share a common horizontal scale of MO signal. Colour coding for each panel is given by the font colour of the labels of the box-and-whisker plots.

Since RMOD provides a quantitative measure of hemozoin content in the blood sample and the detected MO signals are continuous, ROC analysis was conducted to determine the cut-off MO signals corresponding to maximum sensitivity and specificity to detect infection *per se*, as well as infections with specific parasite species. Sensitivity and specificity were determined by selecting the minimum distance from the ROC curve to the [0,1] coordinate indicating 100% sensitivity and specificity. The cut-off values and resulting indicators of agreement between methods are presented in Table II. The ROC curves are included as Supporting Information Fig. S2.

**TABLE II.** **Cut-off values to characterise sensitivity and specificity resulting from ROC analysis per-formed using different reference methods (LM, PCR, RDT) and different infection characteristics.** Here, *κ* is the coefficient of agreement according to Cohen et al.[44] Bold numbers are the means and numbers in parentheses are the 95% conffidence intervals. A table including the predictive values and the area under the ROC curve is provided in the supporting information (Supporting Information Table S2).

| | cut-off (mV/V) | Sensitivity (%) | Specificity (%) | $\kappa$ |
| --- | --- | --- | --- | --- |
| **LM result** |  |  |  |  |
| all LM positive | 4.47 | **0.82** (0.76-0.85) | **0.84** (0.82-0.88) | **0.65** |
| *P. falciparum* | 4.07 | **0.80** (0.74-0.85) | **0.83** (0.78-0.86) | **0.57** |
| *P. vivax* | 5.14 | **0.87** (0.80-0.92) | **0.88** (0.85-0.90) | **0.62** |
| *P.f.+P.v. (mixed)* | 10.23 | **0.95** (0.75-1.00) | **0.95** (0.93-0.97) | **0.54** |
| **PCR result** |  |  |  |  |
| all PCR positive | 3.54 | **0.78** (0.73-0.82) | **0.78** (0.75-0.81) | **0.54** |
| *P. falciparum* | 3.01 | **0.80** (0.74-0.85) | **0.75** (0.71-0.78) | **0.47** |
| *P. vivax* | 4.84 | **0.79** (0.71-0.86) | **0.84** (0.81-0.87) | **0.51** |
| *P.f.+P.v (mixed)* | 5.40 | **0.80** (0.59-0.93) | **0.85** (0.82-0.88) | **0.24** |
| **RDT result** |  |  |  |  |
| all RDT Positive | 4.11 | **0.76** (0.71-0.80) | **0.83** (0.79-0.86) | **0.58** |
| PfHRP2 | 2.58 | **0.65** (0.52-0.77) | **0.70** (0.66-0.73) | **0.16** |
| pLDH | 5.14 | **0.81** (0.71-0.88) | **0.87** (0.84-0.90) | **0.55** |
| PfHRP2+pLDH | 4.11 | **0.81** (0.75-0.86) | **0.83** (0.79-0.86) | **0.57** |

In summary, when based on expert LM as reference standard, ROC analysis indicated that maximum sensitivity and specificity of RMOD for the detection of any *Plasmodium* infection were 82% and 84%, respectively, and the optimal cut-off value to distinguish between infections and non-infections was 4.47 mV/V. The overall agreement between LM and MO was classified as substantial (*κ* = 0.65).[44] Sensitivity and specificity increased for *P. vivax* infections (incl. mixed infections) to 87% and 88%, respectively. When based on PCR as a reference method, MO performed less well with and overall 68% sensitivity and 73% specificity. The overall agreement between PCR and MO was classified as moderate (*κ* = 0.43). When RDT was used as the reference method, sensitivity was 76% and specificity was 83% with an overall agreement classified as moderate-to-substantial (*κ* = 0.58).

One has to keep in mind, that none of the methods used for comparison is a perfect standard, as they also suffer of systematic and stochastic errors. In fact, we will discuss the data obtained for a considerable set of patients later, where RMOD may have detected low-level infections that were not detected by the other methods.

Parasite density as determined by LM correlated well with the MO signals as shown in Fig. 3. On average, *P. falciparum* infected samples resulted in lower MO values as compared to *P. vivax* infected samples with similar parasite density. *P. falciparum* gametocyte containing samples formed a characteristic sub-population, that, on average, exhibited higher MLD values at a similar parasite density when compared with *P. falciparum* asexual stage samples. The correlation of MO values with parasite density was better for *P. vivax* (Spearman Rank *R* = 0.83, *p* < 0.0001) than for *P. falciparum* (Spearman Rank *R* = 0.41, *p* < 0.0001).

**FIG. 3.**
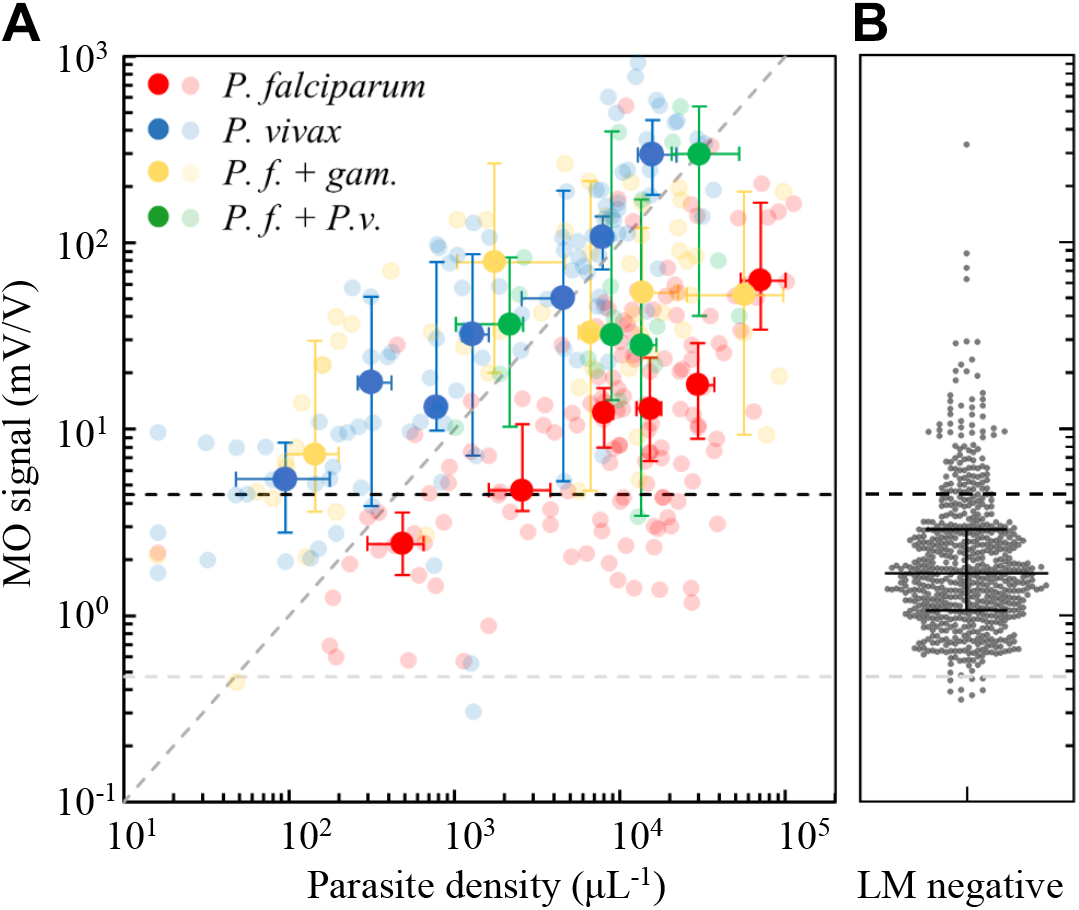
Correlation of the MO diagnostic data with parasite density determined by expert LM. **A** MO signals as function of parasite density for different species, as determined by expert LM. *P. falciparum* mono-infections are represented by red circles, *P. vivax* mono-infections by blue circles and *P.f*.+*P.v*. mixed infections by green circles. Samples with gametocytes in *P. falciparum* mono-infections (*P.f*.+gam.) are represented by orange circles. The larger, solid-coloured circles show median MO signals and median parasite densities for representative parasite density ranges, with 95% CIs as error bars. The smaller, transparent circles show the individual test results. The diagonal dashed line with a slope equal to 1 represents linear proportionality of the MO signal with parasite density. The dashed horizontal black line represents the cut-off value (MO = 4.47 mV/V) determined by the ROC analysis for all LM positive samples (see Table S3). The dashed horizontal grey line is the median MO background signal (0.47 mV/V) determined in malaria naïve volunteers, providing a significantly lower cut-off value. B MO signals of patient samples with undetectable parasites by expert LM. Horizontal lines indicate the median, 1.67 (95% CI: 1.57 *−* 1.78), and the IQR (1.06 *−* 2.89) of the MO signals.

Despite the clear correlation of the MO signal and the parasite density determined by LM, we observed a considerable scatter of the MO signal at a given parasite density. This is natural, as the MO signal depends not only on the density of the parasites but also on the hemozoin content of the parasites, determined by their developmental stage. This alone can explain a considerable variation of the MO signal at any parasite density. However, we also observed considerable MO signal scatter within the sample population which tested negative with all diagnostic methods used for comparison (RDT and LM and PCR), and patients in this group exhibited, on average, higher MO signals than expected from our laboratory-based previous studies.[27]

In addition, the median MO signals in the malaria negative population were statistically significantly higher (more than double) than those measured in a smaller sample of malaria naïve or long-term malaria free volunteers (*n* = 32), as shown in Fig. 4A. Furthermore, within the malaria negative study population, patients who indicated having had malaria in the last two weeks exhibited significantly higher MO signals. In summary, this indicates that recent infection is associated with residual hemozoin levels, impacting MO diagnosis of acute malaria infection. This hypothesis is further substantiated by the observation that MO signals in the malaria negative (by LM and RDT and PCR) sub-population were highly correlated with malaria positivity rate in the overall population (as determined by LM), when both populations were stratified by age (Fig. 4B, Spearman rank correlation coefficient *R* = 0.83; *p* = 0.003). This implies that either i) RMOD detects low-level infections that are not detected by the reference methods or ii) there is a higher residual hemozoin level in populations that are more frequently infected with malaria.

**FIG. 4.**
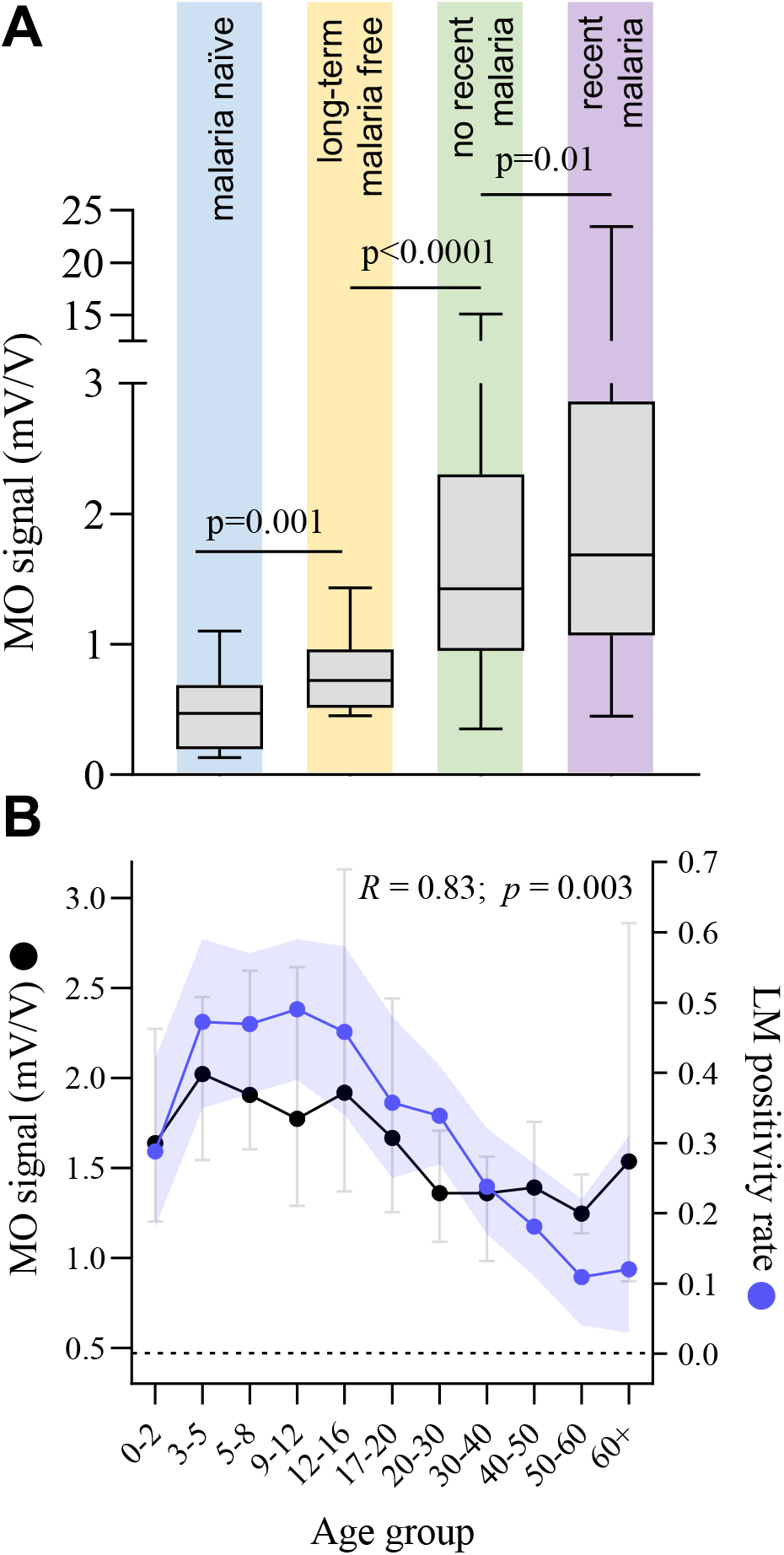
Analysis of MO signals in the population found malaria negative by the reference methods. A Comparison of MO diagnostic data between malaria negative (by LM, PCR and RDT) study participants who reported a recent malaria infection in the two weeks (*n* = 174) preceding the measurement and those who did not report a recent infection(*n* = 254). The graph also shows measurements from *n* = 12 long-term malaria free or malaria naïve volunteers measured during this study as well as previous data from *n* = 20 malaria naïve volunteers measured in non-endemic settings using the same protocol. The distributions were compared using Mann-Whitney U tests resulting in the *p*-values indicated in the graph. B Correlation between LM-based infection positivity rate and MO signals in patients with undetectable malaria in corresponding age groups. LM positivity rate (blue) is shown as proportion with the error band representing the 95% confidence intervals (CI) of proportions. MO signals (black) are shown as medians per age group with error bars representing 95% CI of the medians. The dashed black line corresponds to the median background MO signal in the malaria naïve volunteer population.

## DISCUSSION

Improving malaria diagnosis in support of malaria control and elimination, especially through the development of methods applicable in resource-limited settings is important and timely.[45] Malaria diagnosis is complicated by many factors, including the frequent occurrence of asymptomatic infections with low parasite density.[46]

In this study, we tested a magneto-optical method for automated and rapid measurement of magnetically induced linear dichroism of hemozoin in close to 1000 human blood samples from suspected malaria patients in a high-transmission area of Papua New Guinea (PNG), where both, *P. falciparum* and *P. vivax* are common.[47] In order to evaluate the RMOD performance we conducted a comparison with various conventional diagnostic methods, namely expert LM, RDTs and PCR.

RMOD quantifies the amount of hemozoin in the sample. Correspondingly, we observed a strong correlation between the measured MO signals and parasite density in the peripheral blood samples. This also led to a substantial agreement of RMOD not only with LM but also to moderate-to substantial agreement with RDT and PCR methods. More specifically, when compared to expert LM, RMOD exhibited an 82% sensitivity and 84% specificity to detect any malaria infection, and substantial agreement between RMOD and expert LM, characterized by Cohen’s *κ* = 0.65.[44] Hemozoin-containing trophozoite and schizont stages are known to sequester less frequently in *P. vivax* than in *P. falciparum* infections, resulting in a higher proportion of these stages present in peripheral blood during *P. vivax* infection.[48, 49] Consequently, we observed approximately 10-fold higher average MO signals for *P. vivax* as compared to *P. falciparum* (asexual stages only) at similar parasite densities (compare, e.g., Fig. 2A and Fig. 3). Similarly, for samples containing *P. falciparum* gametocytes, average MO signals were found to be significantly higher in the statistical analyses than for samples containing *P. falciparum* asexual stages only. This is due to the high per-cell hemozoin content found in *P. falciparum* gametocytes.[15, 23, 41]

As a result, RMOD sensitivity and specificity to detect *P. vivax* and mixed infections were significantly higher than that observed for *P. falciparum* when compared to expert LM and PCR, or, when RDT was used as reference method, sensitivity and specificity was observed to be higher in pLDH positive samples as these more frequently contain *P. vivax* (compare Table II). Using expert LM as the main reference method, we found that RMOD detected a higher proportion of *P. vivax* infections than the CareStart RDT (87% vs. 77%) but less *P. falciparum* infections (80% *vs*. 92%).

While MO signals were significantly correlated with parasite density (*P. vivax*: Spearman Rank *R* = 0.83, *p* < 0.0001; *P. falciparum*: Spearman Rank *R* = 0.41, *p* < 0.0001), the scatter of MO signals in the malaria negative population (compare Fig.s 3 and 4) led to considerably higher cut-off levels, below which a measurement is considered negative, than the background MO signal level determined using malaria naïve volunteer samples.[26, 27] This led to a seemingly decreased diagnostic performance. The most plausible hypothesis to explain this observation is, that a considerable proportion of the patients had hemozoin in their peripheral blood at levels still detectable by RMOD, yet in the absence of an infection detectable by any of the methods used for comparison.

Low-density, asymptomatic infections that are undetectable to expert LM, conventional PCR and RDTs are highly prevalent in the study area.[46] Such low-level, undetectable infections may still lead to the accumulation of hemozoin in peripheral blood. In addition, hemozoin persists, in the peripheral circulation, usually inside leukocytes, well beyond the time when all parasites are cleared.[50] *P. falciparum* gametocytes, which, in comparison to asexual stages, contain large amounts of hemozoin, exhibit delayed clearance after antimalarial treatment and may be in circulation for long periods of time.[15, 23, 51] Hemozoin is known to persist in the liver and spleen of previously infected individuals for extended periods of time.[52, 53] On this basis, it is likely that in high transmission settings, such as the setting of the present study, elevated hemozoin levels are maintained in the peripheral blood of a large proportion of the general population, either from concurrent low-level infections that are otherwise undetectable,[54] or from recently treated or cleared infections.

Evidence further supporting this hypothesis, is shown in Fig. 4. Patients indicating a malaria infection within the previous 2 weeks exhibited significantly higher MO signal levels. In addition, samples from *n* = 32 long-term malaria free or malaria naïve volunteers exhibited much lower MO signal values (median: 0.54 mV/V). When malaria negative patients, who had reported recent malaria infection, were excluded from the ROC analysis, overall sensitivity and specificity in comparison to expert LM increased to 86% and 84%, respectively.

In low-transmission and/or elimination settings, which many countries are currently working towards, the residual hemozoin level in the population will be lower, and the MO signal should approach that measured for long-term malaria free and malaria naïve volunteer samples. This will lead to an increase in the ability of RMOD to discriminate infections in these settings. In order to esmalaria the magnitude of improvement, to be expected when lowering the MO signal threshold to the level observed in long-term malaria free and malaria naïve individuals, we used the distribution of MO signals obtained from these samples (*n* = 32) together with the distribution of MO signals from samples that were positive for *P. vivax* by expert LM in the present study (*n* = 124) in a ROC analysis. This resulted in an expected sensitivity of 97% and specificity of 97%, together with a near perfect agreement between LM and RMOD techniques, with *κ* = 0.91.

In conclusion, we present an extensive assessment of a promising novel method to diagnose malaria rapidly and at low cost in a high transmission setting and a population of symptomatic, suspected malaria cases in PNG. The present study shows that in such a setting, the MO diagnostic method performs well in comparison to expert LM-based diagnosis, the most reliable reference method in our study. However, residual hemozoin levels presented a significant limitation, compromising the ability of the method to discriminate between actual and recently cleared infections. This limitation is expected to be reduced in low-transmission settings.

In the current state, RMOD cannot distinguish between parasite species in *P. falciparum* and *P. vivax* coendemic settings. However, RDTs can also not clearly distinguish parasite species. For example, in this study, RDT was able to distinguish *P. vivax* (based on the pLDH-line test result) with 60% sensitivity and 97% specificity. Further development of RMOD is ongoing to address species-specific diagnosis, by taking advantage of the species-specific shape and size distributions of hemozoin crystals.[55]

It is worth considering and further exploring the usefulness of RMOD-based hemozoin detection for malaria diagnosis and in epidemiological studies, in particular in low-transmission and elimination settings. Many countries in these settings experience an increase in the proportion of *P. vivax* infections[56] and it can be expected that *P. vivax* will represent the vast majority of infections in the malaria end-game outside of Africa. Given the rapid and low-cost measurement possible with RMOD and the promising results in particular for *P. vivax*, the method may play a beneficial role in such settings.

Since hemozoin is an intrinsic biomarker of *Plasmodium* spp. infection that persists in peripheral blood (similar to the targets of, e.g., serological antibody tests) it may also qualify RMOD as a tool to detect transmission hotspots in elimination settings.

## MATERIALS AND METHODS

### Study site and sample collection

Madang is located on the North Coast of Papua New Guinea (Fig. 1A). *P. falciparum* and *P. vivax* are highly endemic, and *P. malaria* and *P. ovale* are also present.[43] Capillary blood samples were collected by qualified nurses or community health workers at Yagaum Rural Hospital located outside of Madang and at Madang Town Clinic in 2017–2018.

Blood samples originated from suspected malaria cases presenting as outpatients at the respective clinics. A rapid diagnostic test (RDT; CareStart^™^Malaria, Accessbio Inc., USA) and two blood slides for light microscopy were prepared at enrolment. All RDT positive patients received standard antimalarial treatment according to PNG malaria treatment guidelines. This study received ethical clearance by the PNG Institute of Medical Research (PNGIMR) Institutional Review Board and the PNG Medical Research Advisory Committee (MRAC, #16.45).

Samples from *n* = 12 long-term malaria free and *n* = 20 malaria naïve volunteers were also measured using RMOD and included in the analysis of the present study. All tests were carried out independently and blinded to the results of each other.

### Light microscopy

Light microscopy (LM) was conducted by the PNGIMR microscopy unit consisting of experienced, WHO certified microscopists. Slides were read, and parasite density determined according to WHO guidelines.[57] All slides were read twice, and if there was a significant difference between these first two reads, a third read was conducted by a senior expert microscopist.

### DNA Extraction and Polymerase Chain Reaction

DNA extraction was performed on 200*μ*L of whole blood using DNA extraction kits for 96 well plates (Favorgen Biotech Corp, Taiwan). DNA was eluted in 30*μ*L of RNAse/DNAse free water. The present study employed a two-step realtime polymerase chain reaction (PCR) protocol as described previously using molecular probes targeting Plasmodium 18S rRNA genes. Briefly, a screening PCR to exclude negative samples was first run on all samples.[58] Samples that were positive by the screening PCR were then subjected to a species-specific quantitative real-time PCR (qPCR) as described elsewhere using a CFX96 Touch real-time PCR system (Bio-Rad Laboratories Pty., Ltd., Australia).[59] Therefore, it was possible to quantify parasite density based on gene copy number. For all reactions, 4*μ*L of the DNA eluate was used, corresponding to DNA material from roughly 33*μ*L of the original sample.

### Magneto-optical (MO) measurements

The concept of the rotating-crystal magneto-optical setup and the underlying physical principles of hemozoin detection are described in detail in our former studies[25–27] and the working principle is depicted in Fig. 1 D. In combination with their magnetism, the elongated, rod or brick-shaped hemozoin crystals [55] exhibit anisotropic optical properties, in particular, magnetically induced linear dichroism. Here, we refer to this effect as the magneto-optical (MO) signal, which is the marker-signal of our hemozoin-based diagnosis.[19, 26]

Briefly, the hemolysed blood sample, transferred into a cylindrical sample holder, is inserted into the center of a ring-shaped assembly of permanent magnets, which creates a strong uniform magnetic field (*B* = 1T) at the sample position. This magnetic field induces the coalignment of the hemozoin crystals, and when the magnetic ring is rotated, the co-aligned hemozoin crystals follow this rotation. During the measurement, polarized light from a laser diode is transmitted through the sample in the direction perpendicular to the plane of rotation of the magnetic field. The rotation of the co-aligned dichroic crystals gives rise to a periodic change in the detected intensity. The modulated intensity, divided by the mean intensity, corresponds to the measured ‘MO signal’, which is displayed in the corresponding figures in mV/V units.[25] For the preparation of the blood lysates, 35*μ*L of whole blood was mixed with 315*μ*L of lysis solution(13.3 mM of NaOH and 0.03% v/v of Triton X-100 in distilled water) and allowed to stand for at least 10 minutes to ensure complete lysis. Thereafter, 280*μ*L of the lysate was transferred into the optical sample holders and RMOD were performed without further delay. The optical measurement of a single sample took approximately 3 − 5 minutes (Fig. 1 B). Samples for MO measurements were prepared in triplicate per blood sample.

In general, optical samples were prepared and measured immediately after collection and transport to the laboratory (*n* = 943), but *n* = 21 samples were frozen overnight before preparation and MO measurement due to logistical constraints such as power cuts.

### Data analysis

Data were entered into an electronic data capture system (Epicollect 5, Imperial College). Proportions are presented alongside their exact 95% confidence intervals.[60] Continuous data are presented as means (95% confidence intervals) for normally distributed data and medians with interquartile range or range for not normally distributed data.

Expert light microscopy (LM), PCR and RDT results were used to compare against the RMOD results. Following the recommendation of WHO, LM was used as the gold standard. Methods were compared by calculating the sensitivity and specificity, predictive values and agreement (*κ*) with respect to the reference method.[61] Receiver operating characteristic (ROC) analysis was used to determine the cut-offs for the continuous RMOD data to best predict presence or absence of infection, i.e., to determine sensitivity and specificity by calculating the minimum distance between the ROC curve and the [0,1] coordinate indicating 100% sensitivity and 100% specificity.[62]

## Data Availability

Data will be made available by the corresponding authors upon request

## General

The authors would like to sincerely thank all study participants. We thank Yagaum Hospital and Madang Town Clinic staff for their collaboration in this research study. We would like to thank Professor William Pomat, Dr. Livingstone Tavul, the PNGIMR microscopy unit, molecular parasitology and entomology laboratory staff for their support.

## Author contributions

Conceived the study: IK, SKa; Developed and tested the RMOD prototype, previous volunteer measurements, conducted training: IK, AO, AB, PM, BM; Conducted MO measurements in PNG: LA, CI, PK, TK; Performed PCR analyses: TK, LT, EN; Coordinated Microscopy: LL; Data analysis: AO, SKa, LA, SKr, IK; Wrote first draft: SKa, AO, SKr, IK;

## Coordinated field study

SKa, LJR, ML.

## Data and materials availability

Data will be made available by the corresponding authors upon request.

## SUPPLEMENTARY MATERIALS

Fig. S1. Correlation of parasite density, as determined by LM, and PCR copy number for P. falciparum and P. vivax.

Fig. S2.. Results of the ROC analysis of RMOD versus the conventional diagnostic methods.

Table S1. Diagnostic performance indicators of RDT and PCR methods compared to expert light microscopy as reference standard.

Table S2. Cut-off values to characterise sensitivity and specificity resulting from ROC analysis performed using different reference methods (LM, PCR, RDT) and different infection characteristics.

## Supporting Information

### Magneto-optical diagnosis of symptomatic malaria in Papua New Guinea

**Supporting Information Table S1:** **Diagnostic performance indicators of RDT and PCR methods compared to expert light microscopy as reference standard**. Values are given as percentages and the respective 95% confidence intervals of proportions (in parentheses). Here, PPV, NPV are the positive predictive values and negative predictive values, respectively, and κ is the coefficient of agreement according to Cohen, Landis and Koch [45]. Bold numbers are percentages and numbers in parentheses are 95% confidence intervals of percentages.

| | Sensitivity | Specificity | PPV | NPV | $\kappa$ |
| --- | --- | --- | --- | --- | --- |
| RDT (any infection) | **86.6**<br>(82.4-90.2) | **87.8**<br>(85-90.2) | **78.4**<br>(73.7-82.5) | **92.8**<br>(90.4-94.7) | **0.72**<br>(0.68-0.77) |
| PCR (any infection) | **88.9**<br>(86.2-91.2) | **80.1**<br>(75.3-84.3) | **78.7**<br>(73.8-83.0) | **89.7**<br>(87.1-92.0) | **0.69**<br>(0.64-0.74) |
| *P.f.*-specific PCR* | **78.3**<br>(72.3-83.6) | **92.4**<br>(90.2-94.2) | **75.2**<br>(69.1-80.7) | **93.5**<br>(91.5-95.2) | **0.70**<br>(0.64-0.75) |
| *P.v.*-specific PCR* | **71.8**<br>(63-79.5) | **95.4**<br>(93.8-96.7) | **70.1**<br>(61.3-77.9) | **95.8**<br>(94.2-97.0) | **0.67**<br>(0.59-0.74) |
| mixed infection PCR | **40.0**<br>(19.1-64.0) | **98.2**<br>(97.1-98.9) | **32.0**<br>(15.0-53.5) | **98.7**<br>(97.8-99.3) | **0.34**<br>(0.16-0.52) |

Note: \*includes *P.f./P.v.* detected in mixed infections

**Supporting Information Table S2:** **Cut-off values to characterise sensitivity and specificity resulting from ROC analysis performed using different reference methods (LM, PCR, RDT) and different infection characteristics**. SE, SP, PPV, NPV are the sensitivity, specificity, positive predictive value and negative predictive value, respectively, AUC is the area under the ROC curve, and κ is the coefficient of agreement according to Cohen, Landis and Koch [45]. Bold numbers are the means and numbers in parentheses are the 95% CIs.

| | cut-off | SE (%) | SP (%) | PPV (%) | NPV (%) | AUC | $\kappa$ |
| --- | --- | --- | --- | --- | --- | --- | --- |
| **LM result** |  |  |  |  |  |  |  |
| all LM positive | 4.47 | **0.82**<br>(0.76-0.85) | **0.84**<br>(0.82-0.88) | **0.81**<br>(0.76-0.85) | **0.85**<br>(0.82-0.88) | **0.90**<br>**(0.88-0.93)** | **0.65**<br>(0.59-0.70) |
| *P. falciparum* | 4.07 | **0.80**<br>(0.74-0.85) | **0.83**<br>(0.78-0.86) | **0.62**<br>(0.56-0.67) | **0.92**<br>(0.90-0.94) | **0.88**<br>(0.85-0.91) | **0.57**<br>(0.51-0.63) |
| *P. vivax* | 5.14 | **0.87**<br>(0.80-0.92) | **0.88**<br>(0.85-0.90) | **0.58**<br>(0.50-0.65) | **0.97**<br>(0.95-0.98) | **0.94**<br>(0.91-0.97) | **0.62**<br>(0.55-0.69) |
| *P.f+P.v.* (mixed) | 10.23 | **0.95**<br>(0.75-1.00) | **0.95**<br>(0.93-0.97) | **0.38**<br>(0.24-0.53) | **1.00**<br>(0.99-1.00) | **0.98**<br>(0.96-1.00) | **0.54**<br>(0.40-0.68) |
| **PCR result** |  |  |  |  |  |  |  |
| all PCR positive | 3.54 | **0.78**<br>(0.73-0.82) | **0.78**<br>(0.75-0.81) | **0.65**<br>(0.60-0.70) | **0.87**<br>(0.84-0.90) | **0.84**<br>(0.82-0.87) | **0.54**<br>(0.48-0.5) |
| *P. falciparum* | 3.01 | **0.80**<br>(0.74-0.85) | **0.75**<br>(0.71-0.78) | 0.53<br>(0.48-0.59) | **0.91**<br>(0.88-0.94) | **0.83**<br>(0.80-0.86) | **0.47**<br>(0.41-0.53) |
| *P. vivax* | 4.84 | **0.79**<br>(0.71-0.86) | **0.84**<br>(0.81-0.87) | **0.50**<br>(0.43-0.57) | **0.95**<br>(0.93-0.97) | **0.87**<br>(0.84-0.91) | **0.51**<br>(0.47-0.58) |
| *P.f+P.v* (mixed) | 5.40 | **0.80**<br>(0.59-0.93) | **0.85**<br>(0.82-0.88) | **0.18**<br>(0.11-0.26) | **0.99**<br>(0.98-1.00) | **0.85**<br>(0.75-0.95) | **0.24**<br>(0.15-0.34) |
| **RDT result** |  |  |  |  |  |  |  |
| all RDT Positive | 4.11 | **0.76**<br>(0.71-0.80) | **0.83**<br>(0.79-0.86) | **0.72**<br>(0.67-0.77) | **0.85**<br>(0.82-0.88) | **0.85**<br>(0.82-0.88) | **0.58**<br>(0.52-0.63) |
| PfHRP2 | 2.58 | **0.65**<br>(0.52-0.77) | **0.70**<br>(0.66-0.73) | **0.18**<br>(0.13-0.26) | **0.95**<br>(0.93-0.97) | **0.69**<br>(0.61-0.77) | **0.16**<br>(0.09-0.22) |
| pLDH | 5.14 | **0.81** | **0.87** | **0.51** | **0.97** | **0.87** | **0.55** |
|  |  | (0.71-0.88) | (0.84-0.90) | (0.43-0.59) | (0.95-0.98) | (0.83-0.92) | (0.46-0.63) |
| PfHRP2+pLDH | 4.11 | **0.81** | **0.83** | **0.61** | **0.93** | **0.89** | **0.57** |
|  |  | (0.75-0.86) | (0.79-0.86) | (0.54-0.67) | (0.91-0.95) | (0.86-0.91) | (0.51-0.63) |

**Supporting Information Figure S1:**
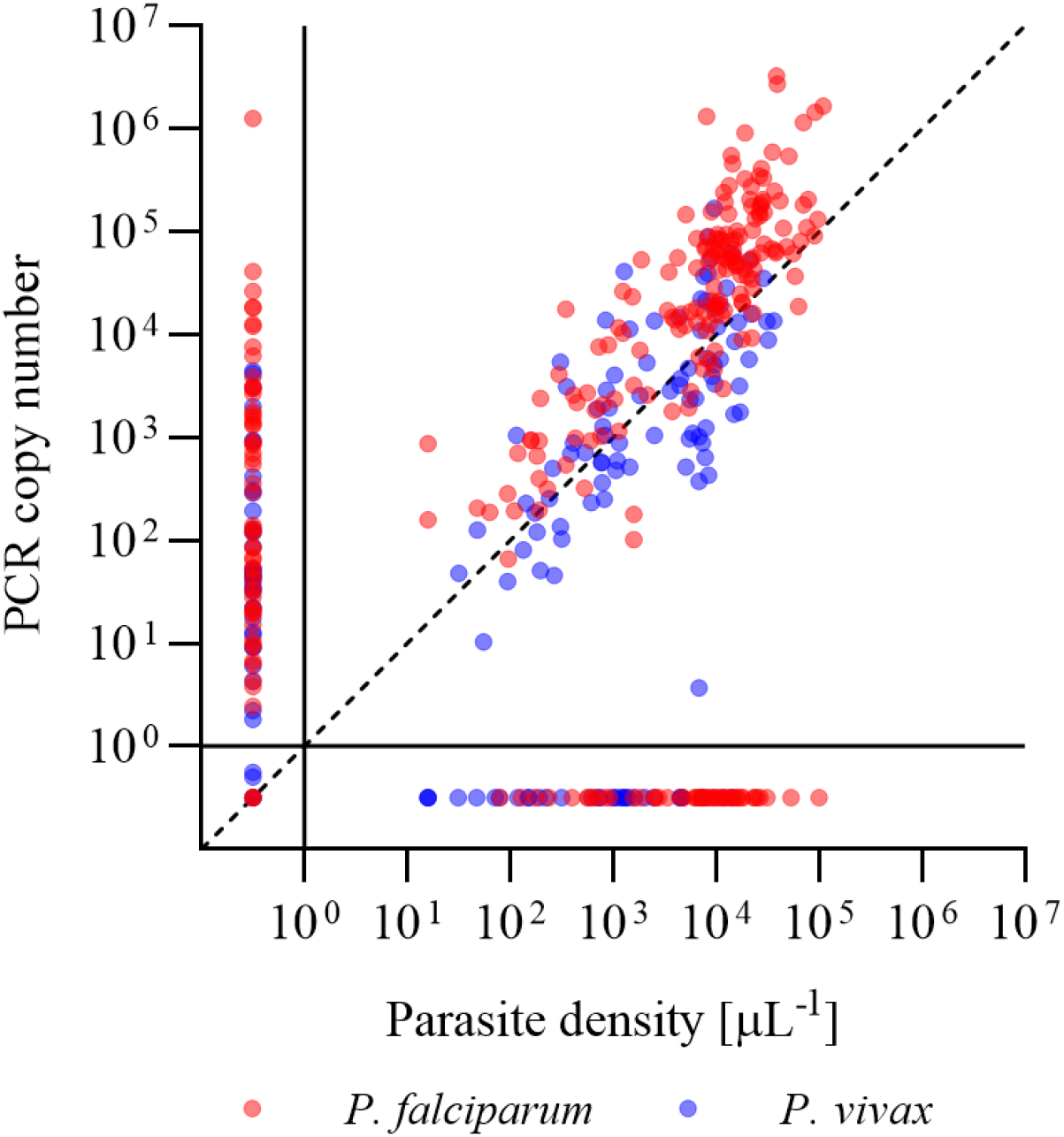
Correlation of parasite density, as determined by LM, and PCR copy number for *P. falciparum* and *P. vivax*. The dashed line represents the line of identity. Symbols displayed at PCR copy <1 and parasite density <1/μL represent PCR and LM negative cases, respectively.

**Supporting Information Figure S2:**
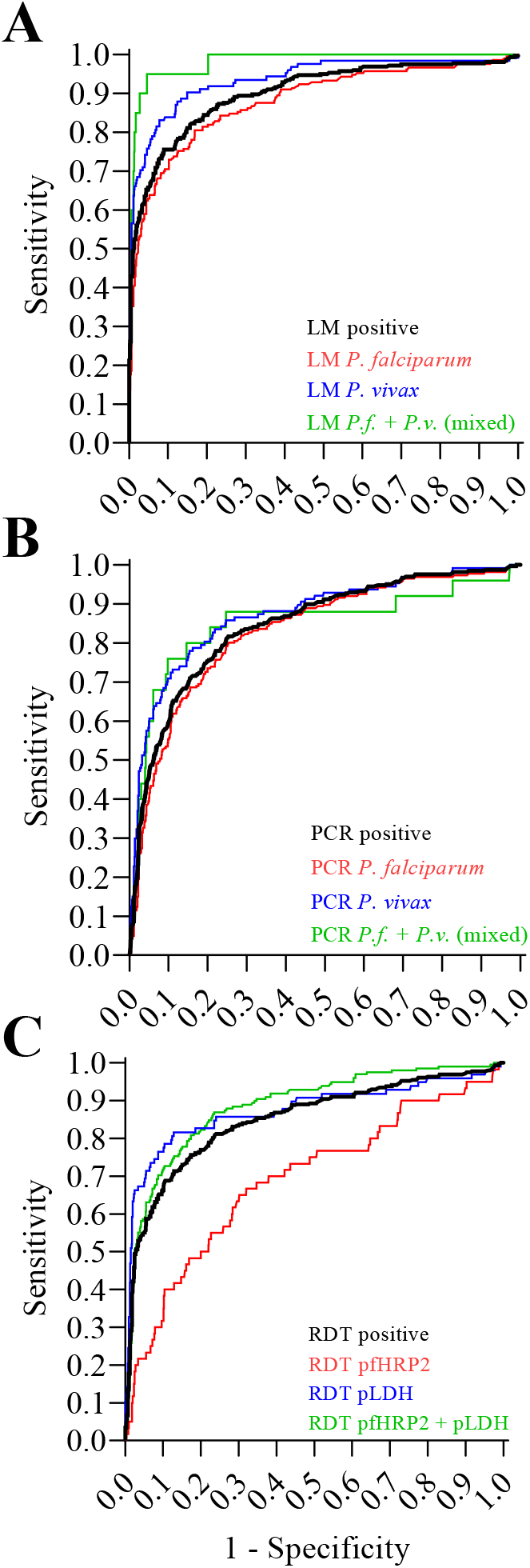
Results of the ROC analysis of RMOD versus the conventional diagnostic methods. Using expert LM as reference method, the red curve represents *P. falciparum* mono-infections, the blue curve represents *P. vivax* mono-infections and the green curve represents *P.f.+P.v*. (mixed) infections. The black curve represents any positive LM result. **Panel B:** Using PCR as the reference method, the red curve represents *P. falciparum* mono-infections, the blue curve represents *P. vivax* mono-infections, the green curve represents *P.f.+P.v*. (mixed) infections as detected by species-specific qPCR. The black curve is the ROC for RMOD versus any species-specific qPCR result. **Panel C:** Using RDT as the reference method, the red curve represents a positive pf*HRP2* line, the blue curve represents a positive pLDH line, the green curve represents measurements on samples where both lines were positive. The black curve represents any positive RDT result. All panels share a common horizontal scale of 1-Specificity.

